# Comparison between 70% ethyl alcohol and 10% formalin as fixative mediums in surgical cooperation campaigns: a pilot study

**DOI:** 10.1101/2024.06.19.24308894

**Authors:** Javier Arredondo Montero, Elena Carracedo Vega, Paula Ortolá Fortes, Mónica Bronte Anaut, Yerani Ruiz de Azúa-Ciria, Adriana Fernández-Ariza, Alejandra Moreno Ibérico, Jessica Paulina Rodríguez, Carlos Bardají Pascual, Rosa Guarch Troyas

## Abstract

**Background:** The lack of adequate resources in international cooperation limits the study of anatomopathological specimens. The literature on potentially inexpensive and available fixation media is scarce.

**Material and methods:** Our surgical team prospectively collected specimens during cooperation campaigns developed in Senegal. Lesions were fixed in parallel in 10% formalin (FF) and 70% ethyl alcohol (AF). Hematoxylin and eosin sections (HE) and immunohistochemistry (IHC) techniques were performed. Images were anonymized and assessed by two senior and two junior pathologists, who evaluated the quality of staining and diagnostic feasibility using an anonymized questionnaire.

**Results:** Three surgical specimens were included: 1 lymph node (3 HE, 4 IHC), one seborrheic keratosis (2 HE, 5 IHC), and one branchial remnant (2 HE, 2 IHC). Fixation times were similar in all the specimens (10-13 days). All FF HE were diagnostic. AF H&E was 100% diagnostic in the 5/7 sections and 75% in the remaining sections. In most cases, pathologists preferred FF. CK7, P40, EMA, CKAE1/AE3, and TTF1 were 100% diagnostic in both groups. CD20, CD45, and EMA were 100% diagnostic (FF) and 75% diagnostic (AF). CD10 was 75% diagnostic (FF) and 25% diagnostic (AF). BCL6 was 75% diagnostic (FF) and 100% diagnostic (AF). IHC preferences were inconsistent.

**Conclusions:** 70% ethyl alcohol has a worse fixation profile than 10% formalin but allows diagnosis in most cases. The immunoreactivity observed is variable depending on the tissue and the stain used. Based on these findings, it can be considered an inexpensive, readily available, and potentially helpful fixation medium for diagnosis in developing countries where surgical cooperation campaigns are conducted. Nevertheless, future studies of larger sample sizes and characterizing other histologic subtypes are needed to confirm these findings.

## Introduction

In surgical pathology, diagnostic accuracy depends on the correct execution of several technical steps, such as specimen collection, fixation, and staining, making fixation one of the process’s first and most important steps. An optimal fixative should be able to prevent putrefaction and autolysis and preserve cellular and tissue details for reliable morphological, immunohistochemical, and molecular studies. Moreover, it should be economic and non-toxic [1-4]. Although suitable quality fixatives are on the market, none is perfect [5,6].

Formalin (aqueous formaldehyde) has been considered the fixative of choice for the last two centuries [6-9]. A 10% solution of formalin (which contains a final 4% concentration of formaldehyde) buffered with phosphate salts to achieve a neutral pH of 7 is usually employed in clinical practice [5-10]. This formalin presentation, usually called neutral buffered formalin (NBF), has significant advantages such as a low cost, a reasonably fast fixation, the possibility of long-term storage, and standardized protocols regarding its use [3,5-7,9]. However, there is extensive evidence regarding its role as a human carcinogen, having been linked to nasopharyngeal cancer and some hematological malignancies such as leukemia [4,5,7,9,11]. In addition, in case of prolonged exposure or accidental contact, it can cause severe mucocutaneous irritation, chemical pneumonitis, and in case of ingestion, potentially life-threatening injuries such as metabolic acidosis, acute renal failure, circulatory shock, and chromosomal alterations [1,3,5]. Due to this, several working groups have proposed to develop new lines of research to find a safer alternative as a histological fixative [4,9].

Another diagnostically relevant aspect is found in the interaction of formalin and specific biological structures: formalin binds DNA, RNA, and proteins (it is a non-coagulant or cross-linking fixative), which potentially can produce a loss of antigenicity and can make molecular biology techniques difficult to interpret [1,5,7-10,12]. Given the significant development of molecular biology in the last decade, this is another critical point that justifies the search for new fixative mediums shortly.

Several types of substances, either commercial or natural, have been proposed and studied as alternatives. The most promising ones are alcohol-based fixatives (ABF) (a mixture of ethanol, methanol, polyethylene glycol, acetic acid, and acetone, among others) [5,7-10]. ABF have shown an excellent fixative profile for morphological and immunohistochemical assessment of pathological specimens. ABF eliminate carcinogenic vapors and are inexpensive, easy to produce, and widely available worldwide. In addition, they offer better DNA, RNA, and protein integrity (they are coagulant fixatives) and more stable specimen preservation-if stored for prolonged periods-when compared to formalin [1,7,10,12]. However, ABF are not free of disadvantages: they are considered highly flammable products, they evaporate quickly, and sometimes they can associate shrinkage of the fixed specimen, which in turn can lead to variability in the stains performed [5,6,9,10,13,14].

Natural fixatives such as honey, sugar, or jaggery have also been considered potential alternative fixatives, especially in areas with scarce economic and material resources (such as low-to-middle-income countries or rural areas) [15]. Of these natural alternatives, bee honey is the one that has shown better results [4]. 10 to 20% dilutions of bee honey have contrasted antibacterial, acidic, and dehydrating properties, and they have demonstrated a similar fixation profile to formalin and alcoholic fixatives, even for nuclear structures [1,3,4,11,14-16].

Of all the options listed above, 70% ethyl alcohol had the best availability profile for our working group. This pilot study aims to assess the methodological feasibility and clinical applicability of using 70% ethyl alcohol as a fixative medium for the pathological study of surgical specimens.

## Methods

### Study design

Our surgical team conducted this prospective, observational study during international cooperation campaigns developed in Velingara, Saint Louis, and Kolda (Senegal). This study included a single group of patients whose biological samples were equally divided into two different fixation media: 1) 10% Formalin (FF) and 2) 70% Ethyl alcohol (AF).

### Sample collection

Biological samples were taken from patients with a lesion that was indicated for surgical resection. The specimens were processed immediately after excision, being sectioned into two fragments of the same size at the expense of their major axis. One of the fragments was placed in a commercial 10% Formalin solution with less than 2.5% methanol (Biopsafe biopsy container, 20 mL, Denmark ©). The other fragment was placed in a commercial solution of 70% ethyl alcohol stained with methylene blue (Alcool Bleu, Valdafrique ©).

### Sample processing

The fixation time was the same for both groups of samples, lasting between 10 and 13 days. The difference in fixation days is because some samples were taken in the first days of the campaign and others in the last days. All samples were processed in parallel. Dehydration and paraffinization of the samples were carried out following a standard protocol. An initial Hematoxylin & eosin from each specimen were evaluated by a consultant pathologist, who oriented the lesions and chose a representative immunohistochemical staining panel. All IHC stainings were performed in parallel on the AF and FF following the standard staining protocols (Roche Diagnostics ©).

Before its evaluation, all sections were digitized (Philips Management System ©). To ensure the study’s rigor, digital photographs were taken at the same magnification level and of the same area for each section to be compared. The images were then anonymized, coded, and pairwise matched (i.e., if the H&E of a piece fixed in FF was designated as 1, the H&E of the same piece fixed in AF was defined as 1B. The assignment of 1 or 1B to AF or FF was made randomly and so forth). The same system was applied to the IHCs.).

## Evaluation forms

Two questionnaires, one for the H&E stainings and one for the IHC stainings, were designed to evaluate the samples.

### Hematoxylin & Eosin evaluation form

In this questionnaire, two questions were asked for each crystal evaluated:

1. Does the staining allow to identify the histological structures adequately (Yes/No)?
2. Which of the two stains is better quality (1 vs. 1B, 2 vs. 2B, 3 vs. 3B, etc…)?

### Immunohistochemical evaluation form

In this questionnaire, three questions were asked for each crystal evaluated:

1. What is the subjective degree of staining? (Artifact/No staining/Weak/Moderate/Strong)
2. Is it adequately distinguished whether the staining is cytoplasmic or nuclear? (Yes/No)
3. Which of the two stains is better quality (1 vs. 1B, 2 vs. 2B, 3 vs. 3B, etc…)?

### Histological evaluation

Two senior pathologists (more than 20 years of clinical experience) and two junior pathologists (less than five years of clinical experience) evaluated the samples. The four participating pathologists were blinded to the surgical specimen, the diagnosis, and the fixative medium used for each section by the abovementioned method. The information from the questionnaires was entered into a database and subsequently analyzed.

### Statistics

For descriptive statistics, percentages were used for categorical variables.

### Research ethics board committee

Local authorities were consulted and confirmed that the health area where the study was conducted did not have a local ethics committee to evaluate the project. The ethical principles of the Declaration of Helsinki (2013) were followed. All patients or their legal representatives signed an informed consent form before their inclusion in the study. All patient data was anonymized following current international legislation.

## Results

Three surgical specimens were included in this study:

1. One lymph node located in the submental region, approximately 1.5 cm of major axis, belonging to a male of African descent (patient’
ss age range: 5-10 years) compatible with atypical lymphoid hyperplasia. This lymph node was resected under the suspicion of a lymphoproliferative process. 3 H&E sections and 4 IHC were performed on it: CD10 (clone SP67), CD20 (clone L26), CD45 (clone RP2/18), BCL6 (clone GI191E/A8).
2. One skin lesion belonging to a male of African descent (patient’
ss age range: 35-40 years), compatible with seborrheic keratosis. 2 H&E sections and 5 IHC techniques were performed on it: CK7 (two sections) (clone SP52), P40 (clone BC28), EMA (two sections) (clone E29).
3. One cervical congenital skin lesion present in a male of African descent (patient’
ss age range: 5-10 years), compatible with a branchial remnant. 2 H&E and 2 IHC were performed on it: CKAE1/AE3 (clone AE1/AE3/PCK26), TTF1 (clone SP141).

Regarding H&E, all FF H&E (7 sections) were diagnostic. AF H&E was considered diagnostic by all pathologists in 5 sections and by 3 of 4 pathologists (2 seniors and one junior) in the two remaining sections. In most cases, pathologists preferred FF. The results obtained from the H&E questionnaire are shown in Table 1.

Regarding IHC staining, CK7, P40, EMA, CKAE1/AE3, and TTF1 were considered diagnostic by all pathologists in both groups. CD20 and CD45 were considered diagnostic by all pathologists in the FF group and by 3 of 4 pathologists in the AF group. In both cases, the same senior Pathologist considered stainings non-diagnostic. CD10 was considered diagnostic by 3 of 4 pathologists in the FF group but only by one pathologist in the AF group. BCL6 was considered diagnostic by 3 of 4 pathologists in the FF group and by all pathologists in the AF group. Regarding artifacts, they were only reported by senior pathologists, mainly in lymphoid tissue sections. IHC preferences were inconsistent. The results obtained from the IHC questionnaire are shown in Table 2. Figure 1 shows some of the most representative sections of the AF and FF groups.

**Fig. 1.**
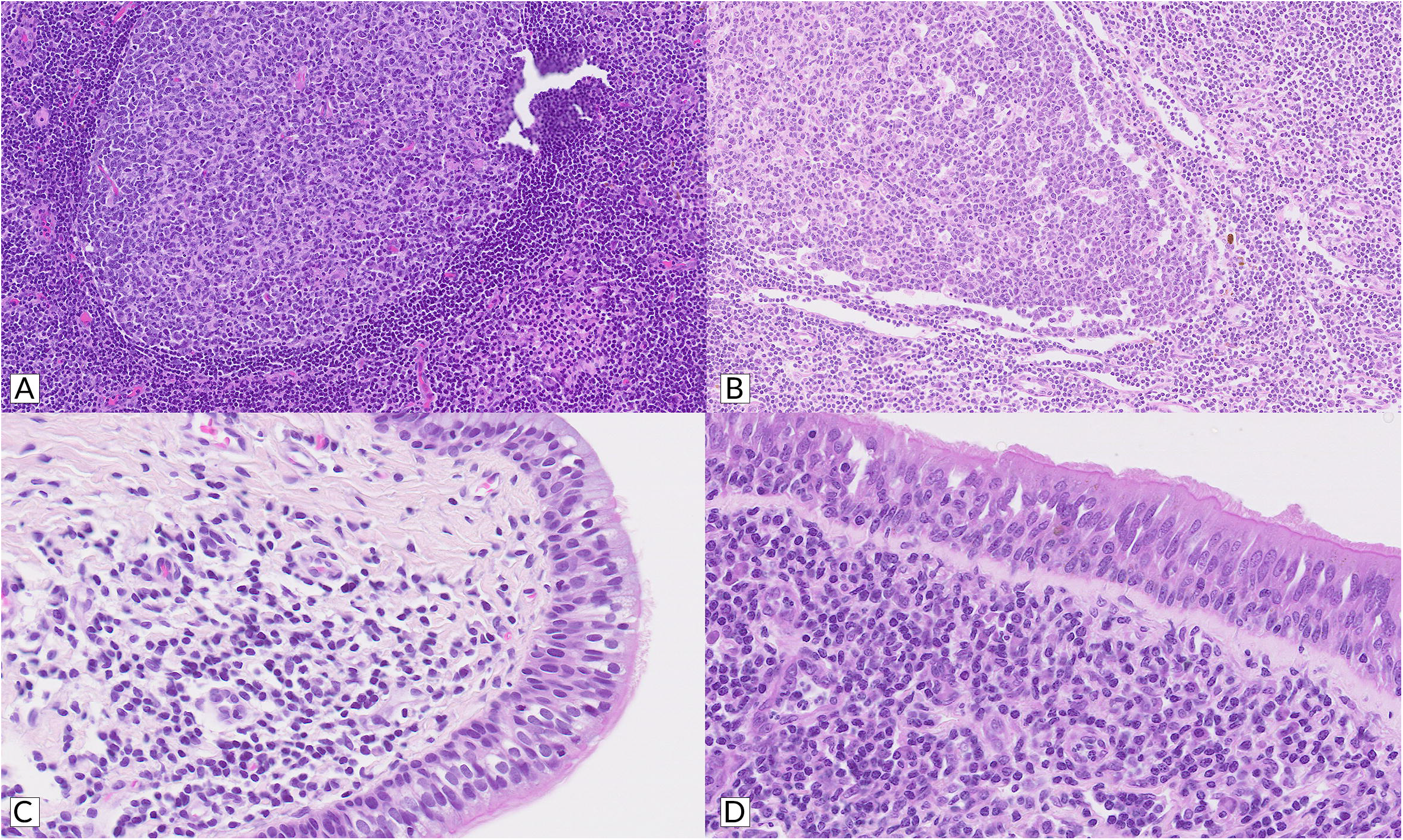
**A, B**: H&E stained sections showing part of a lymph node fixed in formalin 10% (A) and ethyl alcohol 70% (B). **C, D:** H&E stained sections showing a ciliated cylindrical epithelium lining a branchial cyst fixed in formalin 10% (C) and ethyl alcohol 70% (D). Although after alcohol fixation, there is greater eosinophilia and loss of contrast in the tissue, it is possible to distinguish the tissue architecture in both cases.

**Fig. 2.**
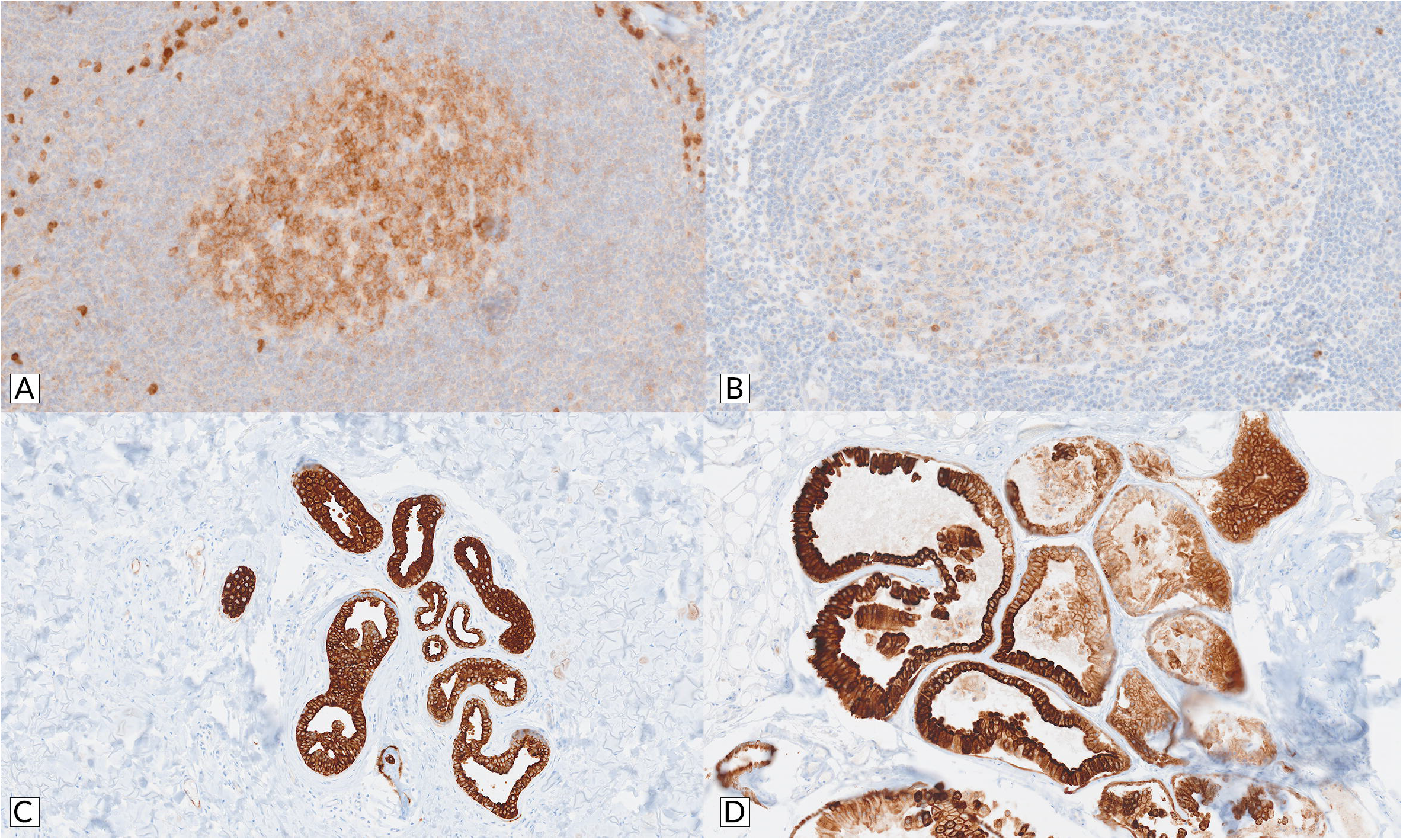
**A, B**: IHC stained sections (CD10) of a lymph node fixed in formalin 10% (A) and ethyl alcohol 70% (B). **C, D:** IHC stained sections (CK7) of a cutaneous lesion fixed in formalin 10% (C) and ethyl alcohol 70% (D). In the case of alcohol fixation, the stains are weaker and patchier, but they still allow for the distinction of morphological features.

## Discussion

The present work constitutes a pilot field test to evaluate the fixative capacity of 70% ethyl alcohol compared to the standardized 10% formalin medium used in clinical practice at our center. This work aims to find economical and available alternatives for the study of anatomopathological specimens obtained in international cooperation surgical campaigns. The preliminary results of this work show that although, in general terms, the quality of fixation (evaluated in terms of final staining results) is inferior to that of 10% formalin, 70% ethyl alcohol has potential as a fixative in international cooperation campaigns since in most sections of our study it allows the diagnosis to be established.

Concerning precedents in the literature, as we stated, there are multiple natural and industrial alternatives to formalin. In our case, the wide availability of 70% ethyl alcohol in local pharmacies led us to opt for this means of fixation. The commercial presentation we obtained was tinted with methylene blue (as a protective measure to avoid accidental ingestion). Still, we do not consider that this had any influence whatsoever on the study we have carried out. We believe it may be interesting to explore alternatives such as ethyl alcohols at different concentrations or honey (considering that there may be problems at the customs level because it is an imported/exported food product).

Regarding the specimens’ fixation time, we consider this a variable that is difficult to control. Surgical campaigns have a variable duration, which, in our case, is usually ten days. This implies that if a sample is collected on the first day of the campaign, it will take at least 13 days for this sample to be processed. If these diagnostic circuits get standardized, patients could be distributed so that those with potentially analyzable lesions could be operated on at the end of the campaign. However, this will not always be feasible. In this regard, we believe it would be optimal to identify the percentage of ethyl alcohol to be applied depending on the expected days of fixation. However, the literature on this issue is limited.

Another relevant aspect of this first stage of specimen processing is the environmental conditions: in Senegal, we generally find temperatures between 35 and 40 degrees Celsius. Although the specimens were protected from sun exposure, the potential impact of external agents on the fixation process must be considered.

It is also pertinent to highlight the medical-legal aspect of the international transport of anatomopathological specimens in international cooperation contexts. This area has been scarcely studied and requires urgent regulation.

Our findings showed better results with H&E than IHC stainings, which is consistent with potential alterations in tissue antigenicity resulting from a suboptimal fixation process. We believe that the poorer results obtained with the lymphoid tissue studied are also consistent with this proposal, given that these tissues are more labile and more susceptible to tissue degradation, which contributes to the fact that they present alterations in their antigenicity more easily.

The specific tissue representations we present constitute a first approximation to the study of 70% ethyl alcohol as a fixation medium. Still, we believe that it is necessary to extend the present study with new tissue representations to better characterize the behaviour of this fixative.

Finally, another interesting point may be to “get used” to seeing pieces fixed in this material. Given that the pathologists included in this study have always worked with pieces fixed in 10% formalin, progressive training in the assessment of pieces fixed in alcoholic media may contribute to improving the diagnostic performance attributable to these fixation media.

The main strengths of this work are its novel design, the extensive characterization of the methodology employed, the use of standardized procedures for sample processing and staining, the use of digitization for section assessment, and the participation of 4 blinded pathologists to evaluate the specimens. Nevertheless, this work has significant limitations, such as the low sample size or lack of representation of all human tissues. Although we believe the questionnaire we have applied has potential usefulness for studies similar to ours, it must be considered that it is not validated and involves an essential component of subjectivity concerning some of its questions (such as the preference between stains).

In conclusion, 70% ethyl alcohol is an inexpensive, safe, and widely available fixation medium that can potentially be used in international surgical cooperation campaigns to study anatomopathological specimens. To validate these findings, more extensive studies with larger sample sizes, greater tissue representation, and rigorous designs that address the limitations of our work are needed.

## Data Availability

All data on this study are available upon justified request through the author in correspondence.

## Acknowledgments

We want to thank the patients who participated in this study. Despite their difficult situation, they enrolled altruistically in this project. Thank you for trying to make the world a better place.

## References

[1] Ozkan N, Salva E, Cakalağaoğlu F, Tüzüner B. Honey as a substitute for formalin? Biotech Histochem. 2012 Feb; 87(2):148–53. doi: 10.3109/10520295.2011.590155. Epub 2011 Aug 23. PMID: 21859382.

[2] Kumarasinghe MP, Constantine SR, Hemamali RL. Methanol as an alternative fixative for cytological smears. Malays J Pathol. 1997 Dec; 19(2):137–40. PMID: 10879255.

[3] Muddana K, Muppala JNK, Dorankula SPR, Maloth AK, Kulkarni PG, Thadudari D. Honey and olive oil as bio-friendly substitutes for formalin and xylene in routine histopathology. Indian J Dent Res. 2017 May-Jun; 28(3):286–290. doi: 10.4103/ijdr.IJDR_246_16. PMID: 28721993.

[4] Yee AWM, Oo PS, Aye SN, Lim WJ, Chee VCX, Krishnappa P. Natural fixatives alternative to formalin in histopathology: A systematic review. Med J Malaysia. 2023 Jan; 78(1):98–108. PMID: 36715199.

[5] Shetty JK, Babu HF, Hosapatna Laxminarayana KP. Histomorphological Assessment of Formalin versus Nonformalin Fixatives in Diagnostic Surgical Pathology. J Lab Physicians. 2020 Dec; 12(4):271–275. doi: 10.1055/s-0040-1722546. Epub 2020 Dec 30. PMID: 33390677; PMCID: PMC7773438.

[6] Ali Jamal A, Abd El-Aziz GS, Hamdy RM, Al-Hayani A, Al-Maghrabi J. The innovative safe fixative for histology, histopathology, and immunohistochemistry techniques: “pilot study using shellac alcoholic solution fixative”. Microsc Res Tech. 2014 May; 77(5):385–93. doi: 10.1002/jemt.22356. Epub 2014 Mar 13. PMID: 24633954.

[7] Zanini C, Gerbaudo E, Ercole E, Vendramin A, Forni M. Evaluation of two commercial and three homemadefixatives for the substitution of formalin: a formaldehyde–free laboratory is possible. Environ Health. 2012 Sep 4; 11:59. doi: 10.1186/1476-069X-11-59. PMID: 22947094; PMCID: PMC3506558.

[8] Benerini Gatta L, Cadei M, Balzarini P, et al. Application of alternative fixatives to formalin in diagnostic pathology. Eur J Histochem. 2012 Jun 29; 56(2):|p e12. doi: 10.4081/ejh.2012.12. PMID: 22688293; PMCID: PMC3428961.

[9] Buesa RJ. Histology without formalin? Ann Diagn Pathol. 2008 Dec; 12(6):387–96. doi: 10.1016/j.anndiagpath.2008.07.004. Epub 2008 Sep 9. PMID: 18995201.

[10] Rahman MA, Sultana N, Ayman U, et al. Alcoholic fixation over formalin fixation: A new, safer option for morphologic and molecular analysis of tissues. Saudi J Biol Sci. 2022 Jan; 29(1):175–182. doi: 10.1016/j.sjbs.2021.08.075. Epub 2021 Aug 27. PMID: 35002406; PMCID: PMC8716893.

[11] Srii R, Peter CD, Haragannavar VC, Shashidara R, Sridhara SU, Srivatsava S. Bee Honey as a Safer Alternative for Routine Formalin Fixation. Kathmandu Univ Med J (KUMJ). 2017 Oct.-Dec.; 15(60):308–312. PMID: 30580347.

[12] Chung JY, Song JS, Ylaya K, et al. Histomorphological and Molecular Assessments of the Fixation Times Comparing Formalin and Ethanol-Based Fixatives. J Histochem Cytochem. 2018 Feb; 66(2):121–135. doi: 10.1369/0022155417741467. Epub 2017 Nov 10. PMID: 29125916; PMCID: PMC5794201.

[13] Bostwick DG, al Annouf N, Choi C. Establishment of the formalin-free surgical pathology laboratory. Utility of an alcohol-based fixative. Arch Pathol Lab Med. 1994 Mar; 118(3):298–302. PMID: 8135636.

[14] Priyadarshi A, Kaur R, Issacs R. Honey as a Cytological Fixative: A Comparative Study With 95% Alcohol. Cureus. 2022 Aug 18; 14(8):e28149. doi: 10.7759/cureus.28149. PMID: 36148184; PMCID: PMC9482673.

[15] Pandiar D, Baranwal HC, Kumar S, Ganesan V, Sonkar PK, Chattopadhyay K. Use of jaggery and honey as adjunctive cytological fixatives to ethanol for oral smears. J Oral Maxillofac Pathol. 2017 May-Aug; 21(2):317. doi: 10.4103/jomfp.JOMFP_224_15. PMID: 28932048; PMCID: PMC5596689.

[16] Singh A, Hunasgi S, Koneru A, Vanishree M, Ramalu S, Manvikar V. Comparison of honey with ethanol as an oral cytological fixative: A pilot study. J Cytol. 2015 Apr-Jun; 32(2):113–7. doi: 10.4103/0970-9371.160563. PMID: 26229248; PMCID: PMC4520042.

