## Supplementary material for "Comparison between 70% ethyl alcohol and 10% formalin as fixative mediums in surgical cooperation campaigns: a pilot study": Table 1

| **Section** | | **Junior Pathologist #1** | **Junior Pathologist #2** | **Senior Pathologist #1** | **Senior Pathologist #2** |
| --- | --- | --- | --- | --- | --- |
| **Lymph node. H&E #1** | | Prefers FF | Prefers FF | Prefers FF | Prefers FF |
|  | AF | AD | AD | AD | AD |
|  | FF | AD | AD | AD | AD |
| **Lymph node. H&E #2** | | Prefers FF | Prefers FF | Prefers FF | Prefers FF |
|  | AF | AD | AD | AD | AD |
|  | FF | AD | AD | AD | AD |
| **Lymph node. H&E #3** | | Prefers FF | Prefers FF | Prefers FF | Prefers FF |
|  | AF | AD | AD | AD | AD |
|  | FF | AD | AD | AD | AD |
| **Skin lesion. H&E #1** | | Prefers FF | Prefers FF | Prefers FF | **Prefers AF** |
|  | AF | **DAD** | AD | AD | AD |
|  | FF | AD | AD | AD | AD |
| **Skin lesion. H&E #2** | | Prefers FF | Prefers FF | Prefers FF | **Prefers AF** |
|  | AF | **DAD** | AD | AD | AD |
|  | FF | AD | AD | AD | AD |
| **Cervical lesion. H&E #1** | | Prefers FF | Prefers FF | Prefers FF | Prefers FF |
|  | AF | AD | AD | AD | AD |
|  | FF | AD | AD | AD | AD |
| **Cervical lesion. H&E #2** | | Prefers FF | Prefers FF | Prefers FF | Prefers FF |
|  | AF | AD | AD | AD | AD |
|  | FF | AD | AD | AD | AD |

**Figure 1. Main Results of the H&E questionnaire.**

**AF:** Ethyl-alcohol 70% fixation; **FF:** Formalin 10% fixation; **AD**: Allows Diagnosis. **DAD**: Doesn’t allow diagnosis
