## Supplementary material for "Comparison between 70% ethyl alcohol and 10% formalin as fixative mediums in surgical cooperation campaigns: a pilot study": Table 2

| **Section** | | **Junior Pathologist #1** | **Junior Pathologist #2** | **Senior Pathologist #1** | **Senior Pathologist #2** |
| --- | --- | --- | --- | --- | --- |
| **Lymph node. CD10** | | Prefers FF | Prefers FF | Prefers FF | Prefers FF |
|  | AF | **Weak staining. DAD** | Weak staining. AD | **Weak staining*. DAD** | **Moderate staining*. DAD** |
|  | FF | Moderate staining. AD | Moderate staining. AD | Strong staining. AD | **Moderate staining*. DAD** |
| **Lymph node. CD20** | | Prefers FF | Prefers FF | Prefers FF | Prefers FF |
|  | AF | Strong staining. AD | Strong staining. AD | Weak staining*. AD | **Strong staining*. DAD** |
|  | FF | Strong staining. AD | Strong staining. AD | Strong staining. AD | Strong staining. AD |
| **Lymph node. CD45** | | Prefers FF | Prefers FF | Prefers FF | Prefers FF |
|  | AF | Strong staining. AD | Strong staining. AD | Weak staining*. AD | **Strong staining*. DAD** |
|  | FF | Strong staining. AD | Strong staining. AD | Strong staining. AD | Moderate staining. AD |
| **Lymph node. BCL6** | | Prefers FF | **Prefers AF** | Prefers FF | **Prefers AF** |
|  | AF | Weak staining. AD | Moderate staining. AD | Weak staining. AD | Weak staining. AD |
|  | FF | Weak staining. AD | Moderate staining. AD | Strong staining. AD | **Weak staining*. DAD** |
| **Skin lesion. CK7** | | **Prefers AF** | **Prefers AF** | NP | **Prefers AF** |
|  | AF | Moderate staining. AD | Strong staining. AD | Strong staining. AD | Strong staining. AD |
|  | FF | Weak staining. AD | Moderate staining. AD | Strong staining. AD | Moderate staining. AD |
| **Skin lesion. CK7** | | Prefers FF | Prefers FF | NP | Prefers FF |
|  | AF | Moderate staining. AD | Strong staining. AD | Strong staining. AD | Strong staining. AD |
|  | FF | Strong staining. AD | Strong staining. AD | Strong staining. AD | Strong staining. AD |
| **Skin lesion. P40** | | Prefers FF | Prefers FF | NP | Prefers FF |
|  | AF | Moderate staining. AD | Strong staining. AD | Moderate staining. AD | Moderate staining. AD |
|  | FF | Moderate staining. AD | Moderate staining. AD | Strong staining. AD | Strong staining. AD |
| **Skin lesion. EMA** | | **Prefers AF** | **Prefers AF** | NP | Prefers FF |
|  | AF | Weak staining. AD | Moderate staining. AD | Strong staining*. AD | Moderate staining. AD |
|  | FF | Weak staining. AD | Moderate staining. AD | Strong staining. AD | Strong staining. AD |
| **Skin lesion. EMA** | | Prefers FF | Prefers FF | NP | Prefers FF |
|  | AF | Moderate staining. AD | Strong staining. AD | Strong staining*. AD | Strong staining. AD |
|  | FF | Moderate staining. AD | Strong staining. AD | Strong staining. AD | Moderate staining. AD |
| **Cervical lesion. CK AE1AE3** | | **Prefers AF** | **Prefers AF** | NP | Prefers FF |
|  | AF | Strong staining. AD | Strong staining. AD | Strong staining. AD | Strong staining. AD |
|  | FF | Strong staining. AD | Strong staining. AD | Strong staining. AD | Strong staining. AD |
| **Cervical lesion. TTF1** | | Prefers FF | Prefers FF | Prefers FF | Prefers FF |
|  | AF | Weak staining. AD | Weak staining. AD | Weak staining. AD | Weak staining. AD |
|  | FF | Moderate staining. AD | Moderate staining. AD | Strong staining. AD | Strong staining. AD |

**Figure 1. Main Results of the IHQ questionnaire.**

**IHQ**: Immunohistochemical staining; **AF:** Ethyl-alcohol 70% fixation; **FF:** Formalin 10% fixation; **AD**: Allows distinction between cytoplasmic or nuclear staining. **DAD**: Doesn’t allow distinction between cytoplasmic or nuclear staining; **NP**: Doesn´t have preferences between both stainings *****: refers artifact
